# Optimising diagnostic yield in highly penetrant genomic disease

**DOI:** 10.1101/2022.07.25.22278008

**Authors:** Caroline F. Wright, Patrick Campbell, Ruth Y. Eberhardt, Stuart Aitken, Daniel Perrett, Simon Brent, Petr Danecek, Eugene J. Gardner, V. Kartik Chundru, Sarah J. Lindsay, Katrina Andrews, Juliet Hampstead, Joanna Kaplanis, Kaitlin E. Samocha, Anna Middleton, Julia Foreman, Rachel J. Hobson, Michael J. Parker, Hilary C. Martin, David R. FitzPatrick, Matthew E. Hurles, Helen V. Firth, the DDD Study

## Abstract

**Background:** Pediatric disorders include a range of highly genetically heterogeneous conditions that are amenable to genome-wide diagnostic approaches. Finding a molecular diagnosis is challenging but can have profound lifelong benefits.

**Methods:** The Deciphering Developmental Disorders (DDD) study recruited >33,500 individuals from families with severe, likely monogenic developmental disorders from 24 regional genetics services around the UK and Ireland. We collected detailed standardised phenotype data and performed whole-exome sequencing and microarray analysis to investigate novel genetic causes. We developed an augmented variant analysis and re-analysis pipeline to maximise sensitivity and specificity, and communicated candidate variants to clinical teams for validation and diagnostic interpretation. We performed multiple regression analyses to evaluate factors affecting the probability of being diagnosed.

**Results:** We reported approximately one candidate variant per parent-offspring trio and 2.5 variants per singleton proband, including both sequence and structural variants. Using clinical and computational approaches to variant classification, we have achieved a diagnosis in at least 34% (4507 probands), of whom 67% have a pathogenic *de novo* mutation. Being recruited as a parent-offspring trio had the largest impact on the chance of being diagnosed (OR=4.70). Probands who were extremely premature (OR=0.39), had *in utero* exposure to antiepileptic medications (OR=0.44), or whose mothers had diabetes (OR=0.52) were less likely to be diagnosed, as were those of African ancestry (OR=0.51).

**Conclusions:** Optimising diagnosis and discovery in highly penetrant genomic disease depends upon ongoing and novel scientific analyses, ethical recruitment and feedback policies, and collaborative clinical-research partnerships.

## INTRODUCTION

Genomic sequencing has made extraordinary strides towards identifying novel molecular causes for rare monogenic disorders, and is becoming increasingly available in diagnostic clinics throughout the world.^1,2^ Pediatrics has particularly benefited from the use of high-throughput next generation sequencing technologies, partly because of the high clinical need and potential for lifelong impact of diagnosis and treatment.^3^ In addition, the early presentation of severe disease makes genetic diagnosis more tractable as causal variants are largely absent from control datasets.^4^

Progress in pediatric rare disease genomics has been spearheaded by numerous diagnostic research groups across the world.^5,6^ One of the first studies to combine large-scale genomic research with individual patient feedback was the Deciphering Developmental Disorders (DDD) study,^7–9^ with >33,500 participants with exome sequence and microarray data and rich clinical phenotypes recorded by >200 clinicians across the UK and Ireland. Here we outline the analytical strategies developed over a decade by the DDD study to identify and classify thousands of new molecular diagnoses, and investigate factors affecting the probability of receiving a diagnosis.

## METHODS

### STUDY OVERVIEW

The DDD study was granted UK Research Ethics Committee (REC) approval by the Cambridge South REC (10/H0305/83) and Republic of Ireland REC (GEN/284/12). A multicentre research collaboration was set up with all 24 Regional Genetics Services, and a management committee (comprising clinicians, scientists and a bioethicist) was created to provide ongoing ethical oversight (**Table 1**). In addition to genomic and data scientists, a social scientist was employed to do ethics research.^10^

**Table 1.** Ethical considerations in the DDD Study. The DDD study depended crucially upon integration of ethics into decision-making and collaboration-building, both upfront and throughout the project, allowing important ethical questions to be identified and ethical policies to be developed through a consensual process.

| **Ethical Domain** | **Key Issues** | **Resolution within DDD Study** |
| --- | --- | --- |
| **Building and maintaining partnerships at clinical-research interface** | • Ensuring trust between researchers, clinicians, and patients/families.<br>• Trade-off between creating large research dataset versus maintaining small clinical cohorts.<br>• Managing practical ethical considerations throughout the lifecycle of the DDD study. | • Scientific scope limited to understanding causes of developmental disorders.<br>• Local training sessions and regular discussion with stakeholders around project planning and decisions.<br>• Regular DDD management committee meetings and annual national collaborators meetings. |
| **Consent, capacity, and eligibility** | • Most DDD probands lack capacity to give consent, either due to young age or intellectual disability.<br>• Initial DDD eligibility limited to those <16 years, creating inequity; however, recruitment of adults lacking capacity is extremely challenging in Scotland.<br>• Confidentiality of DDD study participants should be protected where possible. | • Detailed consent materials and website developed for families/guardians.<br>• New consent materials written, and recruitment opened to adults with/without capacity in England, Wales, Northern Ireland, and Ireland.<br>• Pseudonymised IDs used throughout study; minimum data required for research stored and personal identifiable data (such as date of birth) not stored within DECIPHER. |
| **Sample inclusion, collection, and verification** | • Balance between scientific benefit of sampling parents and clinical concerns around scope and data management.<br>• Many children with developmental disorders are very distressed by hospital visits to have blood samples drawn.<br>• Potential for misattributed parentage and sample mix-ups in DDD (either within families, at recruitment centres or at the Wellcome Sanger Institute). | • Parents recruited into DDD with the agreement that their data would only be used where it is relevant to understanding their child's disorder.<br>• Saliva sample kits used to collect child and parental samples, allowing sample collection at home.<br>• Genetic 'barcodes' for all samples created using 60 SNP-genotyping; individual and family data cross-checked; discordant samples or biologically unrelated parents excluded from further analysis. |
| **Sharing clinically-relevant variants** | • Public opinion about feedback of incidental findings from genomics research largely unknown and unexplored.<br>• Balance between benefits and harms of returning different types of clinically actionable findings.<br>• Pertinent findings (i.e. potentially relevant to the child's developmental disorder) deemed within the scope of research study and clinical testing, where benefits likely to outweigh harms.<br>• Incidental findings deemed outside the scope and expertise of clinicians/researchers, with unclear relevance particularly in children, where harms likely to outweigh benefits. | • Ethics/social science researcher embedded in DDD study to investigate attitudes amongst the public, patients, scientists and health professionals to feedback of incidental findings in genomics.<br>• DDD documentation states that pertinent findings would be reported to clinical teams for communication with families, but not incidental findings.<br>• DDG2P database and variant filtering rules developed to select plausibly pathogenic variants for reporting into linked DECIPHER records; DDG2P genes associated with adult-onset diseases flagged for review.<br>• Pathogenic variants and phenotypes shared openly via DECIPHER once family had been informed. |
| **Sharing genome-wide variants** | • Requests received from DDD parents for genomic data to be returned directly to them.<br>• Access to research data should be prioritised for the hundreds of clinicians and scientists involved in recruitment and management of DDD families.<br>• Research data should be shared widely with external researchers to advance research, but datasets are sensitive as they relate to severely unwell children and consent is limited to understanding causes of DD. | • Requests for individual/family genomic data declined based on concerns about sample identity, lack of resources to provide informatics support, and inability to mitigate against unintended consequences.<br>• Collaborative Analysis Project system created, with research plans reviewed by management committee and data shared by secure file transfer protocol.<br>• Genomic data shared with *bona fide* researchers under managed access via EGA; anonymised variants of potential relevance shared through DECIPHER as research variants. |
| **Managing withdrawal** | • DDD participants are allowed to withdraw from the study at any time, requiring a range of actions to manage samples, data, and associated records.<br>• Previously shared data cannot be withdrawn and may be required to support published findings. | • Upon receiving a withdrawal request, samples are destroyed, unshared data are removed, and individual DECIPHER records are deleted to break any link to the family.<br>• Data in previously published datasets not withdrawn, as stated in consent materials. |

### COHORT

13,610 cases (88% as parent-offspring trios) were ascertained and recruited between April 2011-2015 by consultant clinical geneticists, facilitated by research nurses/genetic counsellors. Families gave informed consent for participation. Eligibility criteria included any of the following: neurodevelopmental disorders; congenital anomalies; abnormal growth parameters (single parameter >4SD or two or more parameters >3SD above the mean); dysmorphic features; unusual behavioural phenotypes; and genetic disorders with a significant impact for which the molecular basis was unknown. The study was initially limited to probands <16 years at the date of recruitment, but this age limit was later removed (except in Scotland). Most probands had previously undergone clinical chromosomal microarray (85%) and/or single gene testing (53%) but remained undiagnosed. Probands were assigned pseudonymised IDs and basic clinical information, quantitative growth data, developmental milestones and Human Phenotype Ontology (HPO)^11^ terms were recorded for all participants via a bespoke standardised interface in DECIPHER.^12^

### GENOMIC ANALYSES

Detailed assay protocols^13,14^ and variant filtering pipelines^7,15^ have been described elsewhere (**Supplementary Information**). Briefly, three independent genomic assays were performed: whole exome sequencing (WES) of complete family trios and singleton probands; exon-array comparative genomic hybridisation (aCGH) of probands; and genome-wide SNP-genotyping of probands. Multiple different algorithms were used to detect and annotate sequence and structural variants (**Figure 1**). *De novo* mutations (DNMs)^16^ and inheritance status of variants in the proband were determined by comparison with parental data. For clinical reporting, we selected high-quality, rare, non-synonymous variants overlapping genes in the Developmental Disorders Gene2Phenotype database (DDG2P)^17^ with appropriate zygosity and inheritance (where available). We augmented this pipeline with additional analyses to find missing likely causal variants, including: mosaic DNMs;^18^ DNMs creating upstream open reading frames;^19^ DNMs affecting splicing;^20^ DNMs of intermediate size;^21^ mobile element insertions;^22^ mosaic chromosomal alterations;^23^ and ClinVar pathogenic/likely pathogenic variants.^24^

**Figure 1.**
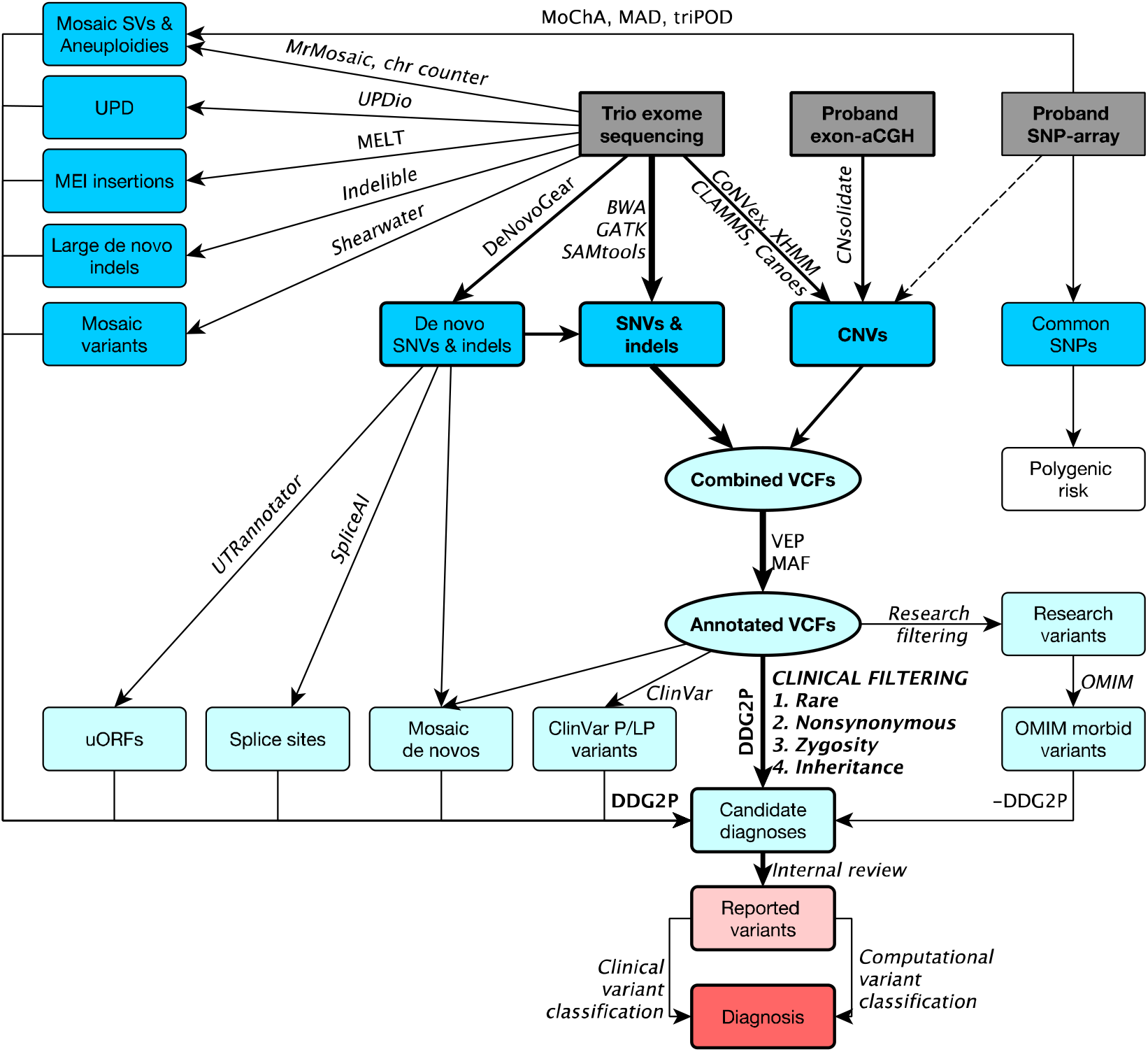
Overview of DDD variant detection and filtering pipelines. Assays are shown in grey boxes, variants in blue boxes, variant subsets in light blue circles, and reported and diagnostic variants in red boxes; variant callers and analytical processes are annotated on arrows (further detail and references in **Supplementary Information**). *aCGH* = array comparative genomic hybridisation; *CNVs* = copy number variants; *DDG2P* = Developmental Disorders Gene2Phenotype database; *indels* = insertions/deletions; *MAF* = minor allele frequency; *MEI* = mobile element insertion; *OMIM* = Online Mendelian Inheritance in Man database; *P/LP* = pathogenic/likely pathogenic (variants in the ClinVar database); *SNP* = single nucleotide polymorphism; *SNVs* = single nucleotide variants; *SVs* = structural variants; *UPD* = uniparental disomy; *uORFs* = upstream open reading frames; *VEP* = Variant Effect Predictor; *VCFs* = variant call files.

### DEFINING A DIAGNOSIS

Candidate diagnostic variants identified bioinformatically were reviewed by a central clinical review panel to evaluate analytical and clinical validity prior to reporting to regional genetics teams via DECIPHER (April 2014-2022, **Figure S1**). The referring clinician then evaluated the reported variant(s), requested diagnostic laboratory confirmation where required, and communicated diagnoses to the family. At the time of writing, clinical classifications of variant pathogenicity (benign/ likely benign/ uncertain/ likely pathogenic/ pathogenic) and contribution to the phenotype (full/ partial/ unknown/ none) were recorded in DECIPHER for 84% of variants. These were supplemented by automated predictions for variant classification criteria (BA1, BS1, BP4, BP7, PVS1, PS1, PS2, PP3 and PM2) based on published guidelines from the American College of Medical Genetics and Genomics (ACMG) and Association of Molecular Pathologists^25^ and UK Association of Clinical Genetic Scientists (ACGS)^26^. A provisional variant classification was calculated using a log-additive Bayesian framework described elsewhere^27^ (**Supplementary Information**). Variants with a posterior probability of >0.9 were classified as likely pathogenic and pathogenic >0.99, or <0.1 as likely benign and benign <0.001. For genes with ≥10 pathogenic/likely pathogenic variants, computational phenotype matching was performed using IMPROVE-DD,^28^ applying the same Bayesian framework to combine variant classifications and gene-disease models; phenotype-based likelihoods were scaled appropriately and used at the evidence equivalent of “Strong”.^27^ Probands were categorised as “diagnosed” if ≥1 variant(s) or ≥2 compound heterozygous variants were annotated as pathogenic/likely pathogenic by either the proband’s referring clinician and/or the predicted classification. Factors influencing the chance of receiving diagnosis (based on clinical annotation only) were investigated using multivariable logistic regression with Bonferroni correction to account for multiple hypothesis testing (**Supplementary Information**).

### DATA AVAILABILITY

Datasets are available under managed access for research into developmental disorders via the European Genome-phenome Archive (EGAS00001000775). Individual pathogenic/likely pathogenic variants are openly accessible with phenotypes via DECIPHER.

## RESULTS

### COHORT CHARACTERISTICS

The DDD study includes 13,450 probands (9,859 in parent-offspring trios) with severe, previously undiagnosed developmental disorders with WES, exon-aCGH and SNP genotyping data, recruited across the UK and Ireland with a median recruitment per centre of 216 probands per million population (range=69-588). The median age was 7 years (range=0-63) at recruitment for probands and 31 years (range=15-90 at the proband’s birth) for parents; 58% of probands were male. A median of 6 HPO terms (range=1-36) were recorded per proband, including 65% with global developmental delay/intellectual disability, and 72% of probands were the only affected member of their family.

### GENETIC FINDINGS

To date, 19,286 potentially pathogenic sequence and structural variants have been identified in DDD probands and reported to referring clinicians through up to six rounds of iterative re-analysis, involving 18 different variant detection algorithms (**Figure 1** and **Table S1**).^7,15^ The majority of variants were identified using a clinically-curated database of 1,840 DD-associated genes (DDG2P)^17^ which was updated at a rate of approximately 100 genes/year through literature curation and cohort-wide enrichment analyses (including 60 novel DD-associated genes identified by DDD; **Figure S1**);^5,13,14,29,30^ 44% of reported variants were in genes added to DDG2P after the first round of reporting in 2014. The majority of reported variants were single nucleotide variants and small insertions/deletions detected using WES data (71% protein-altering, 19% protein-truncating, 3% non-coding variants), while structural variants were identified through a combination of microarray and WES analyses (6% copy number variants, 1% other structural variants; **Figure S2**). On average, one variant was reported per trio versus 2.5 per singleton proband (**Figure S3**), and each new round of analysis resulted in approximately one additional variant being reported for every six trios. Consistent with similar studies,^31^ DNMs in the proband and variants inherited from a mosaic parent (i.e. post-zygotic parental DNMs) in dominant genes provided the highest diagnostic yield, with 79% of reported variants clinically classified as pathogenic/likely pathogenic; in contrast, 32% of variants in autosomal recessive genes, 23% maternally inherited on the X-chromosome, 11% dominantly inherited from an affected parent or with unknown inheritance were clinically classified as pathogenic/likely pathogenic (**Figure 2**).

**Figure 2.**
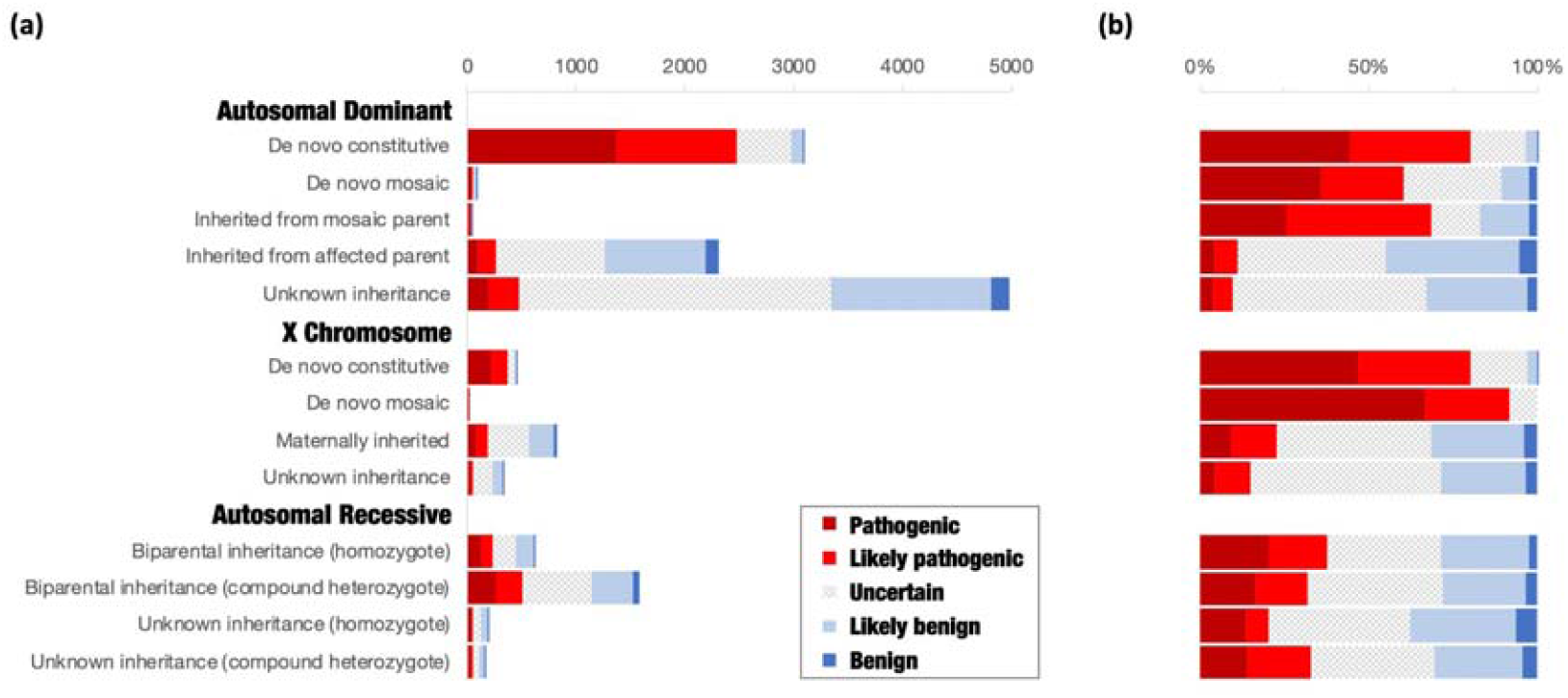
Proportion of clinically classified variants. **(a)** Total number and **(b)** percentage of candidate variants deposited in DECIPHER, separated by mode of inheritance of gene-disease entries in DDG2P and inheritance of variants in probands based on comparison with parental genotypes (trios only).

There was a high rate of concordance between clinical and predicted classifications of variant pathogenicity and benignity (N=4425; sensitivity=99.5%, specificity=85.0%, PPV=96.5%, NPV=97.9%; **Figure S5**).^25–27^ Discrepancies (N=149; 3%) were due to false positive variant calls, incorrect clinical classifications (e.g. atypical disease presentations) or inappropriate ACMG/ACGS criteria assignment (e.g. incorrect disease mechanisms). Based on concordance between clinical and predicted classifications of variant pathogenicity, we estimate that a minimum of 25% of probands (N=3359) are diagnosed, which rises to 32% (N=4237) for predicted only, 34% (N=4507) for clinical only, and 41% (N=5511) for either clinical or predicted (**Figure 3**). Of those who were diagnosed by clinical assertion, 67% (N=3032) have a pathogenic DNM, 12% (N=559) are partially diagnosed, and a further 3% (N=128) have two or more different genetic diagnoses potentially resulting in a composite phenotype.^32^ Although 30% (N=4021) have no reported variants, the rest of the cohort have variants of uncertain significance, of which 0.7% (N=99) have a predicted Bayesian posterior probability of pathogenicity 0.8-0.9. High rates of concordance were also seen in the subset of variants for which we were able to derive a phenotype-based gene-disease model using IMPROVE-DD,^28^ and a further 18 variants of uncertain significance were predicted to be pathogenic/likely pathogenic based on the individual’s phenotype.

**Figure 3.**
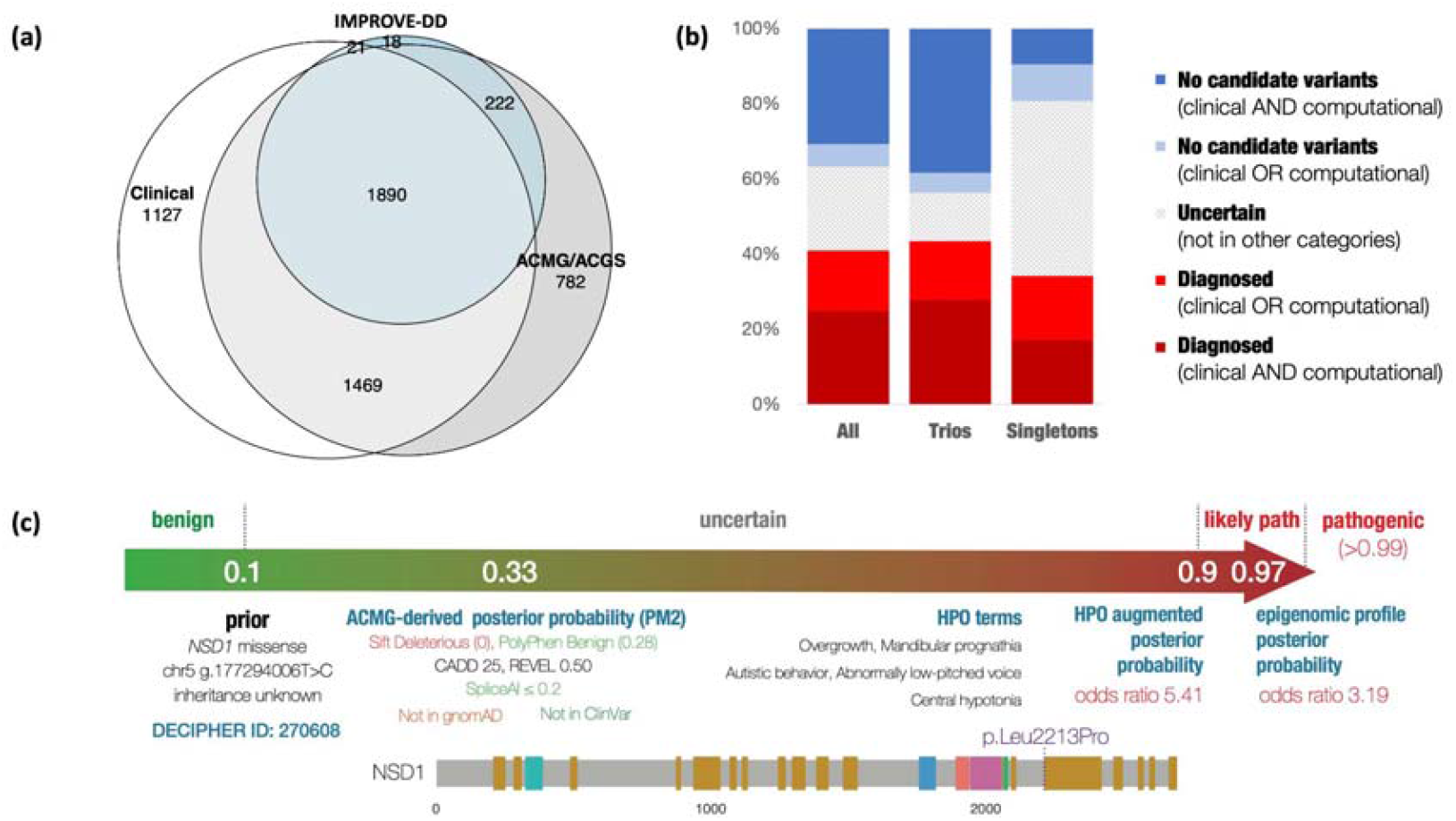
Summary of diagnoses in the DDD study. **(a)** Venn diagram showing overlap of diagnoses based on clinical assertion (white) versus predicted ACMG/ACGS variant classifications (grey), augmented with phenotype-based IMPROVE-DD gene-disease models (blue); figure created using eulerr. **(b)** Diagnostic ranges in trios and singleton probands, based on clinical and/or predicted variant classifications. **(c)** Example of computational Bayesian variant classification, incorporating genotypic and phenotypic data in a DDD proband: only PM2 could be applied to the missense variant, resulting in an uncertain classification, but the proband’s phenotype was consistent with the IMPROVE-DD model for *NSD1*, allowing the variant to be upgraded to likely pathogenic; additional data (e.g. epigenomic profiling)^40^ was used to further increase the robustness of the diagnosis of Sotos syndrome.

### FACTORS INFLUENCING DIAGNOSTIC RATE

We performed multiple logistic regression to investigate how demographic, clinical, phenotypic, prenatal and ancestral factors affected the chance of receiving a clinical diagnosis from the DDD study (**Figure 4**). The model explained ∼14% of the variance. Being in a parent-offspring trio had the largest impact on the chance of being diagnosed (OR: 4.70, 95% CI: 4.16-5.31). Other factors significantly increasing the chance of diagnosis included: having severe intellectual/ developmental delay (OR: 2.41, 95% CI: 2.10-2.76); time since recruitment (increased odds of diagnosis: 1.25 per additional year, 95% CI: 1.20-1.30); being the only affected family member (OR: 1.74, 95% CI: 1.57-1.92) or having fewer affected first-degree relatives (**Figure S6**); having features suggestive of a syndrome (OR: 1.23, 95% CI: 1.12-1.34); and having more organ systems affected (increased odds of diagnosis: 1.08 per additional organ system, 95% CI: 1.06-1.11). Probands born prematurely (OR: 0.73, 95% CI: 0.64-0.82), or who had *in utero* exposure to antiepileptic medications (OR: 0.44, 95% CI: 0.29-0.67) or whose mothers had diabetes (OR: 0.52, 95% CI: 0.41-0.67) were less likely to have a genetic diagnosis. Male sex (OR: 0.72, 95% CI: 0.67-0.79) also reduced the odds of getting a diagnosis, as did increasing homozygosity due to consanguinity (decreased odds of diagnosis: 0.72 for each increase equivalent to the offspring of first cousins, 95% CI: 0.62-0.83). Probands with African ancestry had a lower diagnostic rate than those with other ancestries (OR: 0.51, 95% CI: 0.31-0.78), which was driven by fewer diagnoses in singleton probands (**Figure S7**). Probands with non-European ancestry had more variants reported back to clinical teams (Wilcoxon *p*-value <0.001), particularly in singleton cases (**Figure S8**).

**Figure 4.**
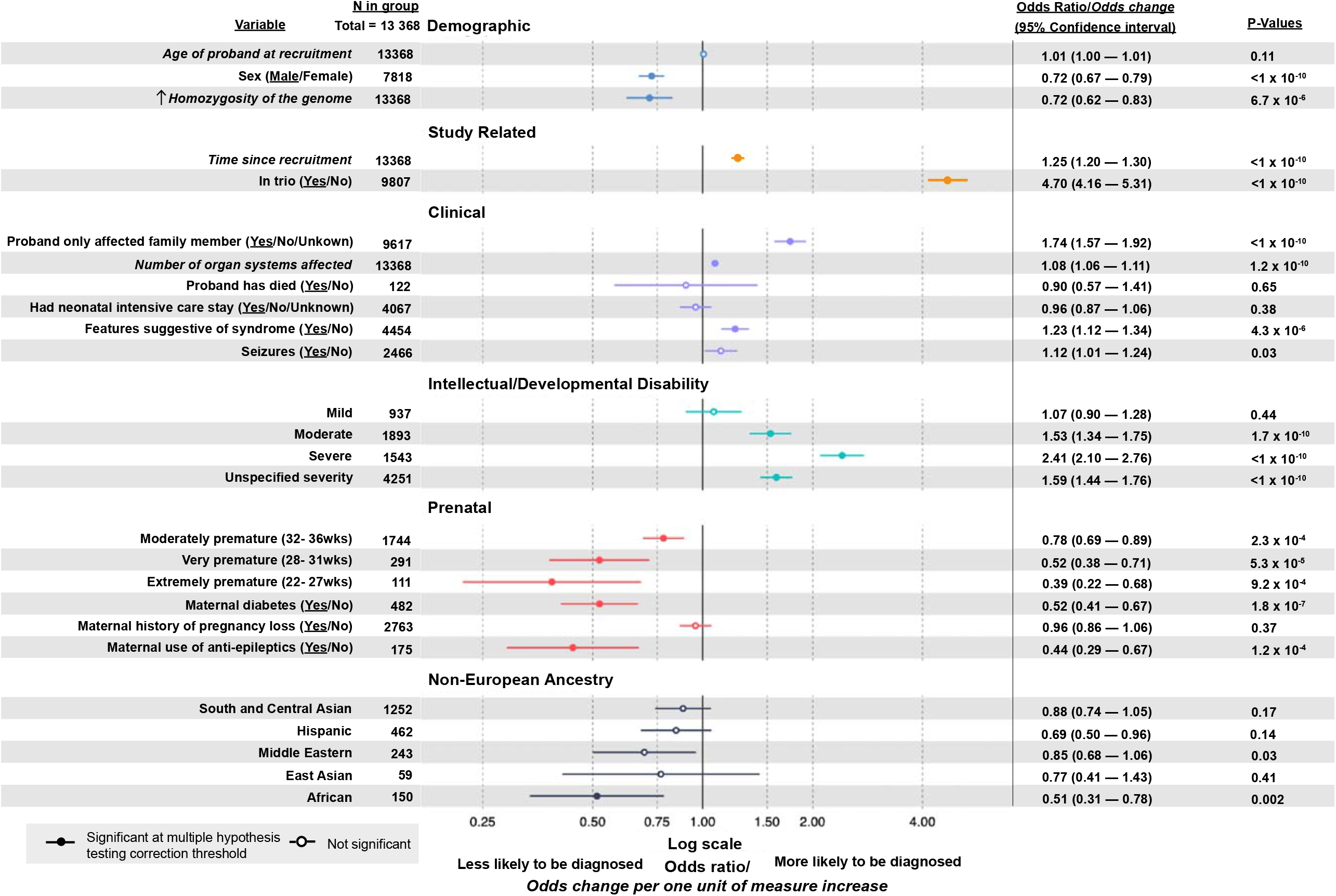
Factors influencing the probability of being diagnosed. Odds associated with being fully or partially diagnosed by the DDD study (based on clinician assertions of variant pathogenicity) are shown for covariates included in a multivariable logistic regression, adjusted for recruitment centre and number of variants reported in DECIPHER. Odds ratios are presented for binary and categorical variables. For quantitative variables (italics), odds change per one unit of measure of increase are presented. P-values and 95% confidence intervals are also shown; filled circle = significant after adjusting for multiple hypothesis testing (Bonferroni p-value<0.003); unfilled circle = not significant; underline in variable column = outcome variable plotted, with N referring to the number of probands in this group. See **Supplementary Information** for further analysis of the number of affected first-degree relatives (**Figure S6**) and ancestry (**Figure S7 and S8**).

## DISCUSSION

The DDD study has identified and communicated molecular diagnoses to thousands of families across the UK and Ireland affected by severe, previously undiagnosed developmental disorders. Our analysis highlights the value of using diverse variant detection algorithms combined with stringent variant filtering rules and iterative variant analysis and classification.^15^ The high burden of pathogenic *de novo* mutations and current diagnostic yield estimate of 34-41% are consistent with similar studies,^33^ but also reflect the challenges of making a robust genetic diagnosis in genetically and phenotypically heterogeneous conditions when both genomic knowledge and disease phenotypes change over time.

Our analysis re-enforces clinical intuition about the likelihood of establishing a molecular diagnosis in developmental disorders (e.g. availability of parental genotype data, as well as sex, ethnicity and phenotypic severity) and moves towards quantifying the expectation of making such a diagnosis. The work also highlights groups with lower diagnostic rates in our cohort (e.g. singletons, families with multiple affected members, and those with non-European ancestry or high consanguinity). Excluding cohort-specific factors, our multivariable logistic regression model predicts that probands in the top decile of probability of being diagnosed have a diagnostic rate of 52% versus 16% in the bottom decile. We hypothesise that the lower diagnostic rate found in probands with certain prenatal factors reflects a larger role for environmental influences in these individuals. Prematurity^34^, maternal diabetes^35^ and *in utero* exposure to antiepileptic medications^36^ are known risk factors for developmental disorders. Further exploration is needed to better understand the relative contributions and interplay of genetic/environmental influences in this cohort.

The genetic architecture of developmental disorders is heterogeneous; although the large burden of highly-penetrant DNMs facilitates both diagnosis and large-scale gene-disease discovery,^5^ the number of composite and partial diagnoses suggests that many individuals are likely to have multiple contributing factors, including rare and common incompletely penetrant genetic variants and non-genetic causes. Under a liability threshold model of disease,^37^ probands who have a significant environmental contribution may require less severe or even no large-effect genetic variants to develop a neurodevelopmental disorder. Nonetheless, statistical burden analyses suggest that many more diagnoses remain in protein-coding genes than in non-coding elements,^38^ which will likely be identified through novel DD-associated gene discovery (especially for dominant disorders), evaluation of incompletely penetrant variants, and functional assays to improve interpretation of existing candidate variants.

The DDD study pioneered a hybrid clinical-research approach, requiring development of new methodologies to facilitate both large-scale analysis and individual variant feedback, which has since become standard practice in genomic medicine. The study primarily recruited infants and children and hence pioneered a conservative approach to individual variant feedback that focussed on diagnosis,^7–9^ whilst exploring attitudes to communicating incidental findings^39^ that influenced subsequent approaches.^1^ A large network of expert clinician-researchers and the integration of ethics at a high level throughout the project lifecycle served to both facilitate collaboration and enable real-time ethical issues to be openly and responsibly addressed (**Table 1**). To date, in addition to making thousands of new diagnoses for patients, the DDD study has resulted in >270 publications (https://www.ddduk.org/publications.html), identified around 60 new disorders and enabled >350 genotype/phenotype-specific projects led by clinician-researchers across all 24 recruitment sites. DECIPHER^12^ was another key component of the DDD study, enabling nationwide recruitment, systematic phenotyping, individual feedback, variant interpretation and data sharing. DECIPHER is a live online platform enabling reported variants to be re-evaluated with current data (e.g. gene-disease associations, population frequencies, co-located variants reported in ClinVar, DECIPHER or publications) each time a patient is reviewed in the clinic, thus facilitating new opportunities for diagnosis as knowledge grows.

Although many of our conclusions are widely applicable across a range of rare diseases, the generalisability is limited by a number of factors. Recruitment of families following routine diagnostic testing (karyotyping, aCGH and targeted single gene testing) resulted in a cohort depleted of clinically recognisable syndromes and large structural variants, reducing the diagnostic yield relative to first-tier testing and skewing the factors affecting getting a diagnosis. The diagnostic yield in DDD therefore represents a conservative estimate with higher yields anticipated if genomic sequencing had been offered as a first-line investigation. Our genotyping approach (WES and microarrays) did not assay most non-coding variants and could not detect all complex structural variants or tissue-specific mosaicism, and our analytical approach was insensitive to incomplete penetrance. Furthermore, the study was not funded to capture longitudinal phenotype data, evaluate parental phenotypes in detail, record the impact of diagnosis on subsequent clinical management of families, or assess social or environmental contributions to developmental disorders – all of which, in retrospect, would have enhanced the project. Finally, despite the large cohort size, due to the enormous genetic and phenotypic heterogeneity, we often had insufficient probands (particularly across ethnicities) with the same ultra-rare condition to enable confident variant interpretation, highlighting the need to aggregate phenotype information and structured electronic health data across cohorts internationally to improve variant interpretation.

## CONCLUSION

The DDD study pioneered nationwide genomic analysis of a large clinical cohort using a hybrid clinical-research model, with the aim of advancing understanding of the genetic architecture of developmental disorders and catalysing improved diagnosis. Converting genomic data into robust clinicomolecular diagnoses for patients requires sophisticated informatics and multidisciplinary expertise. The DDD study shows how the fusion of clinical expertise, genomic science, bioinformatics and embedded ethics can drive diagnosis and discovery in genomic medicine.

## Data Availability

All data produced are available upon request from the European Genome-phenome Archive.

https://ega-archive.org/studies/EGAS00001000775

## ACKNOWLEDGEMENTS

The authors are deeply indebted to the patients and families involved in the DDD study, as well as the UK NHS and Irish HSE genetics services and the many scientists who have worked on DDD data at the Wellcome Sanger Institute (**Supplementary Information**). The DDD study presents independent research commissioned by the Health Innovation Challenge Fund [grant number HICF-1009-003], a parallel funding partnership between the Wellcome Trust and the Department of Health, and the Wellcome Sanger Institute [grant number WT098051]. The views expressed in this publication are those of the author(s) and not necessarily those of the Wellcome Trust or the Department of Health. The study has UK Research Ethics Committee approval (10/H0305/83, granted by the Cambridge South REC, and GEN/284/12 granted by the Republic of Ireland REC). The research team acknowledges the support of the National Institute for Health Research, through the Comprehensive Clinical Research Network. The study uses DECIPHER (https://www.deciphergenomics.org), which is funded by Wellcome [grant number WT206194]. PC was supported by an NIHR Academic Clinical Fellowship.

## Notes

### Competing Interest Statement

The authors have declared no competing interest.

### Funding Statement

This study was funded by the Health Innovation Challenge Fund [grant number HICF-1009-003], a parallel funding partnership between the Wellcome Trust and the Department of Health, and the Wellcome Sanger Institute [grant number WT098051].

### Author Declarations

The Cambridge South Research Ethics Committee of the UK Research Ethics Committees gave ethical approval for this work.

