## Supplementary Information for "Optimising diagnostic yield in highly penetrant genomic disease"


**Full list of DDD scientists and NHS collaborators** 2
- Wellcome Genome Campus 2
- National Health Service/Health Service Executive Regional Genetics Services 2

**Variant detection, annotation and filtering** 4

**Automated ACMG/ACGS variant classifications** 6

**Factors affecting getting a diagnosis** 8

**Supplementary Figures** 11
- Figure S1. Changes in DDG2P and variants reported with time. 11
- Figure S2. Candidate diagnostic variants deposited in DECIPHER. 12
- Figure S3. Number of reported variants per proband. 13
- Figure S4. CADD versus REVEL scores for high impact de novo variants. 14
- Figure S5. Concordance between calculated Bayesian posterior probability of variant pathogenicity and clinical classifications. 15
- Figure S6. Affected status of first-degree relatives alters diagnostic rate. 16
- Figure S7. Comparison of the odds of diagnosis for non-European ancestry probands between trio and singleton cases. 17
- Figure S8. Breakdown of variants reported via DECIPHER across ancestry groupings. 18

**Supplementary Tables** 19
- Table S1. Proportion of reported variants per variant detection algorithm. 19
- Table S2. Covariates included in the multivariable logistic regression. 20
- Table S3. Trio information across different ancestry groupings. 23

**Supplementary References** 24

### Full list of DDD scientists and NHS collaborators

**Wellcome Genome Campus**

Nadia Akawi, Saeed Al-Turki, Kirsty Ambridge, Katrina Andrews, Jana Awada, Daniel Barrett, Jeffrey Barrett, Tanya Bayzetinova, A. Paul Bevan, Eugene Bragin, Simon Brent, Patrick Campbell, Nigel Carter (former PI), Eleni Chatzimichali, V. Kartik Chundru, Stephen Clayton, Panayiotis Constantinou, Manuel Corpas, Fiona Cunningham, Petr Danecek, Ruth Eberhardt, Helen Firth (CI), Tomas Fitzgerald, David FitzPatrick, Julia Foreman, Giuseppe Gallone, Eugene Gardener, Sebastian Gerety, Susan Gribble, Juliet Handsaker, Rachel Hobson, Sarah Hunt, Matthew Hurles (PI), Benjamin Hutton, Phil Jones, Wendy Jones, Joanna Kaplanis, Rosemary Kelsell, Dan King, Netra Krishnappa, Sarah Lindsay, Jenny Lord, Tom Marchant, Hilary Martin, Jeremy McRae, Anna Middleton, Ray Miller, Kate Morely, Matt Neville, Mari Niemi, Michael Parker, Daniel Perrett, Elena Prigmore, Lizze Radford, Di Rajan, Kaitlin Samocha, Hassan Shakeel, Patrick Short, Alejandro Sifrim, G. Jawahar Swaminathan, Anja Thormann, Adrian Tivey, Margriet van Kogelenberg, Parthiban Vijayarangakannan, Nicola Whiffin, Emilie Wigdor, Caroline Wright.

*Supported by core staff in the sample management, sequencing, genotyping and human genetics informatics teams*

**National Health Service/Health Service Executive Regional Genetics Services**

**Referring clinical geneticists**
***Aberdeen:*** Moira Blyth, John Dean (PI), Zosia Miedzybrodzka, Lisa Kier Robertson, Alison Ross.
***Belfast:*** Tabib Dabir, Deirdre Donnelly, Vivienne McConnell, Alex McGee, Shane McKee (PI), Patrick Morrison, Gillian Rea, Fiona Stewart.
***Birmingham:*** Helen Kate Brittain, Louise Brueton, Trevor Cole (former PI), Nicola Cooper, Helen Cox, Farah Fanani, Alison Foster, Rebecca Igbokwe, Lily Islam, Joanna Jarvis, Esther Kinning, Derek Lim, Jenny Morton (former PI), Swati Naik, Mary O'Driscoll, Kai-Ren Ong, Deborah Osio, Corrina Powell, Nicola Ragge, Suresh Somarathi, Hannah Titheradge (PI), Julie Vogt, Denise Williams.
***Bristol:*** Ana Beleza, Lucy Bownass, Simon Bodek, Alan Donaldson, Karen Low, Ruth Newbury-Ecob (PI), Andrew Norman, Ingrid Scurr, Sarah Smithson (PI), Madeleine Tooley.
***Cambridge:*** Ruth Armstrong, Kate Baker, Jenny Carmichael, Helen Firth (PI; DDG2P curation), Simon Holden, Sarju Mehta, Soo-Mi Park, Joan Paterson, Lucy Raymond, Evan Reid, Richard Sandford, Marc Tischkowitz, Geoff Woods.
***Cardiff*:** Ayesha Ahmed, Angus Clarke, Hector Conti, Andrew Fry, Jennifer Gardner, Vani Jain, Arveen Kamath, Oliver Murch, Caroline Pottinger (PI), Annie Procter, Julian Sampson, Francis Sansbury, Ian Tully, Vinod Varghese.
***Dublin:*** Lisa Bradley, Andrew Green, Sally Ann Lynch (PI).
***Dundee:*** Jonathan Berg (PI), David Goudie, Catherine McWilliam.
***Edinburgh:*** David FitzPatrick (former PI; DDG2P curation), Andrew Jackson, Wayne Lam, Anne Lampe (PI), Michael Yates (DDG2P curation).

***Exeter:*** Emma Baple, Carole Brewer, Bruce Castle, Ruth Cleaver, Emma Kivuva (PI), Henrietta Lefroy, Julia Rankin, Charles Shaw-Smith, Peter Turnpenny, Claire Turner, Anna Znaczko.
***Glasgow:*** Panayiotis Constantinou, Rosemarie Davidson, Carol Gardiner, Mark Hamilton, Shelagh Joss (PI), Cheryl Longman, Ruth McGowan, Victoria Murday, Daniela Pilz (former PI in Cardiff), Edward Tobias, John Tolmie, Margo Whiteford.
***Leeds:*** Christopher Bennett, Jennifer Campbell, Angus Dobbie, Verity Harthill, Emma Hobson, Rosalyn Jewell, Alison Kraus, Katrina Prescott (PI), Eamonn Sheridan, Jenny Thomson.
***Leicester:*** Pradeep Vasudevan (PI), Jennifer Hague.
***Liverpool:*** Natalie Canham (former PI in North West Thames), Ian Ellis, Lynn Greenhalgh, Jenny Higgs, Emma McCann, Victoria Mckay, Astrid Weber (PI).
***London (Great Ormond Street Hospital):*** Tazeen Ashraf, Angela Barnicoat, Maria Bitner-Glindicz, Emma Clement, Francesca Faravelli, Jane Hurst (former PI), Wendy Jones (PI), Eleanor Hay, V.K.Ajith Kumar, Melissa Lees, Alison Male, Elisabeth Rosser, Mina Ryten, Richard Scott, Shereen Tadros, Emma Wakeling, Louise Wilson.
***London (Guy's and St Thomas'):*** Fiona Connell, Frances Flinter, Muriel Holder, Melita Irving, Louise Izatt, Dragana Josifova, Shehla Mohammed (PI), Leema Robert, Deborah Ruddy, Mina Ryten.
***London (North West Thames):*** Birgitta Bernhard, Angela Brady, Virginia Clowes, Jan Cobben, Alice Gardham, Neeti Ghali, Susan Holder, Rita Ibitoye (PI), Jessica Anne Radley.
***London (St George's):*** Frances Elmslie, Tessa Homfray, Nayana Lahiri, Sahar Mansour (PI), Meriel McEntagart, Anand Saggar, Kate Tatton-Brown.
***Manchester:*** Siddharth Banka, Catherine Breen, Tracy Briggs, Emma Burkitt-Wright, Kate Chandler, Jill Clayton-Smith (former PI), Charu Deshpande, Dian Donnai, Sofia Douzgou, Elizabeth Jones, Bronwyn Kerr, Kay Metcalfe (PI), Audrey Smith, Helen Stuart.
***Newcastle:*** Marta Bertoli (PI), John Burn, Richard Fisher, Alex Henderson, Tara Montgomery, Ruth Richardson, Miranda Splitt (former PI), Volker Straub, Michael Wright, Laura Yates.
***Nottingham:*** Abhijit Dixit, Jacqueline Eason (PI), Rachel Harrison, Gabriella Jones, Nora Shannon, Ajoy Sarkar, Claire Searle, Mohnish Suri.
***Oxford:*** Edward Blair, Deirdre Cilliers, Andrew Douglas, Richard Gibbons, Usha Kini (PI), Victoria Nesbit, Andrea Nemeth, Joanna Poulton, Susan Price, Debbie Shears, Helen Stewart, Andrew Wilkie.
***Sheffield:*** Meena Balasubramanian, Diana Johnson, Oliver Quarrell, Mick Parker (PI), Alison Stewart.
***Southampton/Wessex:*** Diana Baralle, Nicola Foulds, Victoria Harrison, David Hunt, Daniela Iancu,Mira Kharbanda, Katherine Lachlan, Catherine Mercer, Lucy Side, Karen Temple (PI), Diana Wellesley.

*Supported by NHS scientists in the molecular and cytogenetics diagnostic laboratories involved in variant validation and interpretation, as well as genetic counsellors, research nurses and numerous trainees and administrators involved in patient recruitment and family counselling.*

### Variant detection, annotation and filtering

Saliva samples were collected (Oragene DNA collection kits, DNA Genotek) and DNA extracted (QIAsymphony, Qiagen); blood-extracted DNA was also collected from NHS laboratories for probands where available. Three genome-wide assays were used:

- whole exome sequencing (WES) of the ~2% of the genome that codes for proteins was performed for all probands and parent-offspring trios using Agilent SureSelect (Human All-Exon V3 or V5 Plus with custom ELID C0338371) and Illumina HiSeq;[1,2]
- exon-array comparative genomic hybridisation (aCGH) was performed for all probands using custom 2x 1M probe Agilent microarrays (Amadid No.s 031220/031221); and
- genome-wide SNP-genotyping was performed for all probands using Illumina HumanCoreExome or HumanOmniExpress chips.

Different algorithms were used to detect sequence and structural variants from WES and microarray data (see **Figure 1** in main paper and **Table S1** for relative contributions to reported variants), including:

- BWA,[3] GATK[4] and SAMtools[5] for detection of SNVs and indels;
- DeNovoGear[6] for detection of likely *de novo* SNVs and indels;
- Shearwater[7] for detection of low-level mosaic sequence variants;
- Indelible[8] for detection of medium-sized insertions/deletions;
- MELT[9] for detection of mobile element insertions;
- XHMM,[10] CLAMMS[11] and Canoes[12] as well as two in-house algorithms (CNsolidate and CoNVex) for detection of CNVs from arrays and WES;
- UPDio[13] for detection of uniparental disomy; and
- MoCha,[14] MAD,[15] MrMosaic,[16] triPOD[17] and an in-house chromosome counting algorithm (chr counter) for detection of mosaic structural variants and aneuploidies.

All data were aligned to GRCh37. Variants were combined into single-sample VCFs and annotated using the Ensembl Variant Effect Predictor[18]. Variants were also annotated with minor allele frequencies (MAF) from the 1000 genomes project[19] and Exome Aggregation Consortium[20] as well as internal MAFs derived from unaffected DDD parents. CNVs were annotated with MAFs derived from 1000 UK blood donors and 500 family trios from the Generation Scotland Study analysed on the same 2x 1M probe microarrays.[21] Candidate *de novo* SNVs/indels were additionally annotated with UTRannotator[22] for detection of uORF-altering 5’UTR variants, and SpliceAI[23] for detection of non-canonical splice variants. DeNovoWEST[24] was used for burden analyses of *de novo* variants. A subset of *de novo* variants were orthogonally validated using capillary sequencing.

Candidate diagnostic variants were selected bioinformatically using to the following stringent variant filtering rules (https://github.com/jeremymcrae/clinical-filter/tree/master/clinicalfilter):[25]

- rare (MAF ≤0.0005 for monoallelic, MAF ≤0.005 for biallelic, and MAF≤0.0005 for X-linked modes of inheritance); and
- non-synonymous (stop-gain, frameshift, splice acceptor/donor, missense, in-frame insertion/deletion, transcript ablation/amplification) variants; overlapping

- DDG2P[26] (protein-coding transcript with most severe consequence flagged; where variant consequences on >2 transcripts were equally severe, the canonical or longest transcript was chosen in preference); with
- appropriate zygosity (including monoallelic, biallelic and X-linked modes); and
- appropriate inheritance where available (including *de novo*, autosomal dominant, autosomal recessive, X-linked dominant and X-linked recessive).

Variants were reviewed prior to deposition into pseudonymised DECIPHER records. Variant quality was evaluated using IGV[27] and/or an in-house data visualisation tool (developed for CNVs), and low quality variants were excluded. A subset of variants were evaluated for phenotypic fit by the central review team, focusing particularly on variants in DDG2P genes associated with both child and adult-onset diseases; variants in individuals without a phenotype consistent with published literature were excluded. In addition to variant filtering for clinical reporting purposes, a stringently filtered set of non-synonymous DNMs and recessively inherited likely loss-of-function variants in non-DDG2P and non-OMIM morbid genes[28] in undiagnosed probands were made openly available for research via DECIPHER (https://www.deciphergenomics.org/ddd/research-variants); those variants in OMIM morbid genes were evaluated for phenotypic fit by the central review team, and variants in individuals with a phenotype consistent with published literature were reported back to clinical teams via DECIPHER.

### Automated ACMG/ACGS variant classifications

Predicted variant classification criteria were based on guidelines[29] produced by the American College of Medical Genetics and Genomics (ACMG) with the Association for Molecular Pathology, and updated[30] by the UK Association of Clinical Genomic Scientists (ACGS). Where appropriate, automated application of criteria were predicted based on the most recent published guidelines using the following thresholds:

| RULE | DECIPHER Automated Implementation |
|---|---|
| **PVS1** | Applied for stop-gain, frameshift, splice acceptor/donor variants in biologically relevant transcripts[31] where DDG2P mechanism is "absent gene product" |
| **PS1** | Applied for ClinVar[32] pathogenic/likely pathogenic 2/3/4-star variants |
| **PS2** | Applied for *de novo* mutations (*de novo* status and family relatedness within DDD are confirmed by trio WES) |
| **PM2** | Applied where gnomAD[33] population MAF=0 (for non-biallelic DDG2P genes) or popmax MAF<0.02% (for biallelic DDG2P genes) and coverage >10X |
| **PP3** | Applied for missense variants where CADD[34]≥30 or REVEL[35]≥0.7 or SpliceAI[23]≥0.7 |
| **BA1** | Applied where gnomAD[33] popmax MAF>5% and coverage >10X |
| **BS1** | Applied where gnomAD[33] popmax MAF<5% AND MAF≥0.5% (for non-biallelic DDG2P genes) or MAF≥1% (for biallelic DDG2P genes) and coverage >10X |
| **BP4** | Applied for missense variants where CADD[34]≤10 or REVEL[35]≤0.2 and SpliceAI[23]≤0.2 |
| **BP7** | Applied for synonymous variants where SpliceAI[23]≤0.2 and PhyloP[36]<0.1 |

Conservative evidence-based thresholds for PP3/BP4 were determined by plotting CADD[34] and REVEL[35] scores for *de novo* mutations identified in 31,058 DD trios[24] filtered to significantly enrich for high impact variants, i.e. absent from gnomAD and functional predicted consequence in a relevant DDG2P gene (**Figure S4**). Evidence strengths were combined using a log-additive Bayesian framework outlined in Tavtigian *et al.*,[37] where prior probabilities were set to 0.1 for all variants, and posterior probabilities were calculated using published likelihood ratios for very strong (PVS1, BA1), strong (PS1, PS2, BS1), moderate (PM2) and supporting evidence (PP3, BP4, BP7). Concordance between the Bayesian posterior probabilities and clinical assertions is shown in **Figure S5**.

The following criteria were not automatically applied or used in Bayesian calculations: PS3 (functional studies not possible to automate); PS4 (prevalence and penetrance in controls unknown for most DDG2P genes); PM1 (mutational hotspots impossible to automate currently across all DDG2P genes); PM3 (heterozygous variants in biallelic genes are only reported in DDD probands in the presence of a second allele in trans); PM4 (further work required to automate repetitive regions across all DDG2P genes); PM5 (different amino acid at same site very rarely used in clinic); PM6 (not relevant for DDD, see PS2); PP1 (co-segregation information not available within DDD); PP2 (further work required to determine appropriate thresholds and appropriate DDG2P mechanisms); PP4 (DDG2P includes highly heterogeneous conditions); PP5 (no longer recommended by ClinGen);[38] BS2 (prevalence and penetrance in controls unknown for most DDG2P genes); BS3 (functional studies not possible to automate); BS4 (co-segregation information not available within DDD); BP1 (missense variants known to cause loss-of-function); BP2 (heterozygous variants in biallelic genes are only reported in DDD probands in the presence of a second allele in trans); BP3 (further work required to automate repetitive regions across all DDG2P genes); BP5 (alternative diagnoses usually unknown at the time of assessment); and BP6 (no longer recommended by ClinGen).[38]

### Factors affecting getting a diagnosis

**PCA ancestry classification**
The ancestry classifications were assigned using principal component analysis (PCA) and UMAP. Data from the 1000 Genomes project and the Human Genome Diversity Project (HGDP) were used as a reference, with the DDD data projected onto the reference principal components (PCs). We retained only variants common to all three datasets. The reference data were filtered for MAF (<1%) and genotype missingness (<90%). Long-range linkage disequilibrium (LD) regions were removed. Following this, the data were merged with the DDD data (also filtered for genotype missingness <90%) and LD pruning was performed using a LD $R^2$=0.2 (the plink flag used was --indep-pairwise 50 5 0.2), leaving 23,507 SNPs. PCA was done on the reference data using the smartpca-fastmode from the EIGENSOFT software, and the DDD data were projected onto those PCs. The first 7 PCs were used by UMAP, run using the uwot R package using the default parameters, to create distinct continental clusters. These were used to assign the ancestry labels to the DDD individuals.

**Determining genome-wide homozygosity ($F_{ROH}$) as a proxy for consanguinity**
Regions of homozygosity (ROHs) were called from the exome data using bcftools/roh using the following parameters: bcftools roh -b -e all -G30 -I -a 6.6e-05 -H 5e-06 -m MAPFILE, where MAPFILE is a genetic map file derived from the 1000 Genomes phase 3 data. For each individual, the total length of ROHs (in base pairs) was summed up and then divided by $3x10^9$ to determine the approximate fraction of the genome in ROHs ($F_{ROH}$ ).

**Methods of multivariable regression analysis**
We used the data gathered from each participant via their standardised recruitment form, together with their genetic findings, to investigate factors influencing the chance of an individual receiving a diagnosis using multivariable logistic regression. A proband was classified as diagnosed based on previously described criteria. The covariates included in the model described in **Table S2**. A total of 82 probands were excluded from the model (<1% of total dataset) due to missing information, or suspected errors in the phenotyping (e.g. implausibly low reported gestational age). Bonferroni correction was performed to account for multiple hypothesis testing (p<0.05/number of covariates, i.e. 0.05/19 = 0.003). We later added an interaction term between ancestry and trio status into the model and ran the model separately in trios and singleton cases. All statistical analyses were performed with R software, version 3.6.1 (R Foundation for Statistical Computing).

**Supplementary discussion note on results of multivariable regression analysis**
The results of the multivariable regression model provide an insight and quantification of factors that influence an individual's chance of being diagnosed within DDD. The effect sizes of factors found to alter diagnostic rate are likely to be influenced by a mix of actual underlying biology of developmental disorders, our current knowledge of the genetic aetiology of development disorders, and DDD design, recruitment and analysis methods. Further details are outlined in **Table S2**.

**Demographic factors**

We found that age of proband at recruitment had no effect on diagnostic rate. It should be noted that recruitment of DDD probands occurred following clinical testing (at a point when microarrays became widely available as first-line clinical tests for DD but whole exome/genome sequencing was not yet available clinically), rather than reflecting when the probands initially presented with developmental disorders. The lower diagnostic rate in males compared to females in DDD was expected. A large body of evidence supports that females require a higher burden of genetic variation before presenting with neurodevelopmental disorders.[39-41] Pathogenic variants in females are therefore more likely to be easier to identify and assess due to being, generally, more severe.[39]

**Study-related factors**

Being in a trio had the largest impact on the chance of diagnosis among study related factors. A trio design has previously been shown to increase diagnostic rates in developmental disorders,[25,40] particularly due to its ability to identify *de novo* mutations. Notably, however, a recent meta-analysis of exome sequencing for neurodevelopmental disorders did not find that having more family members sequenced improved diagnostic rates, although the heterogeneity of study types and cohorts included may have confounded these analyses.[41] Probands recruited to DDD earlier had higher diagnostic rates. This may be partly because probands recruited earlier in the study were enriched for phenotypes more tractable to genetic diagnosis, but another factor is that clinicians have simply had more time to review relevant their genetic variants and decide which are pathogenic.

**Clinical factors**

A number of clinical factors affected diagnostic rate. Firstly, probands with more severe phenotypes had a higher chance of diagnosis: both increasing number of organ systems affected and more severe intellectual disability were associated with higher diagnostic rates. Akin to the higher diagnostic rate in females,[42,43] this probably reflects that more severe phenotypes are likely to have more damaging genetic mutations on average. Thus, their pathogenic variants are easier to identify and assess compared to milder cases which may possibly have an oligogenic or polygenic rather than monogenic basis. Probands with features suggestive of a syndrome also had higher diagnostic rate, perhaps because genotype-phenotype correlation and variant assessment are easier in the presence of distinctive syndromic features.

We expected that probands who had died may also have generally more severe variants and therefore have higher diagnostic rates; however, the number of individuals in the study who have died since recruitment is small and we did not find any significant differences in diagnostic rates. Whether the proband had a neonatal intensive care stay, a seizure phenotype, or history of pregnancy loss also had no significant effect on diagnostic rate after multiple testing correction.

Probands who were the only affected member of their family had higher diagnostic rates. Breaking this down further, we found that the more close relatives are affected, the lower the diagnostic rate for probands in DDD (**Figure S6**). This may reflect ascertainment bias. Probands in pedigrees with multiple affected individuals are more likely to have been considered for inclusion in other next generation sequencing studies at the time, perhaps meaning that such pedigrees in DDD are depleted of cases easily tractable to genetic diagnosis. It is possible that some families with multiple affected individuals do not have a simple monogenic cause but rather an oligogenic cause or simply high polygenic load.[44] It could in part reflect bioinformatic filtering difficulties too: if the parent is also affected and a dominant cause is assumed, the search space of possibly pathogenic variants is very large.[25]

**Ancestry**

Probands with African ancestry had a low diagnostic rate compared to those with European ancestry, which was driven by singleton cases being particularly difficult to diagnose. The lack of ancestry-matched control cohorts to estimate allele frequency is one probable cause: we find that both fewer variants are filtered out in the clinical pipeline for non-European probands and the variants deposited in DECIPHER for clinicians for review appear to be more difficult to interpret (**Figure S7 and S8**). A trio design seems to partially counteract this, probably through the ability to filter out inherited rare variants and highlight high-probability pathogenic *de novos*. Unfortunately, probands with non-European (especially African) ancestry are less likely to have been sequenced as a trio, compounding inequality in diagnostic rates between ancestries (**Table S3**). Another potential issue contributing to the lower diagnostic rate is that developmental disorder phenotypes are generally less well defined in patients of African ancestry, particularly features of facial dysmorphism.[45] This can make clinical interpretation of variants more challenging, perhaps leading to more reported variants being rated ‘uncertain’ or remaining unclassified. These findings support ongoing and future work to curate larger databases of allele frequencies in African-ancestry cohorts and dysmorphology resources for African ancestry individuals to improve diagnostic rate,[45–47] as well as encouraging any interventions that could help to increase trio uptake in African-ancestry populations.

### Supplementary Figures

#### Figure S1. Changes in DDG2P and variants reported with time.

Gene-disease entities were added to the DDG2P database following curation of the literature by consultant clinical geneticists or burden analyses within the DDD study. Entries include an assessment of the evidence-level (e.g. number of independent families identified), mode of inheritance (i.e. monoallelic, biallelic, X-linked), mechanism of pathogenicity (i.e. loss-of-function, missense/inframe) and associated phenotypes.
Participants were sequenced and analysed in batches based on recruitment date, sample receipt and family trio status. Variant filtering was repeated over the course of the study to enable evaluation of novel variants and variants in newly included genes. As a result of this iterative variant filtering strategy, some probands were evaluated up to six times and all were evaluated at least twice. Following evaluation, variants were deposited into DECIPHER in batches for evaluation by clinical teams.

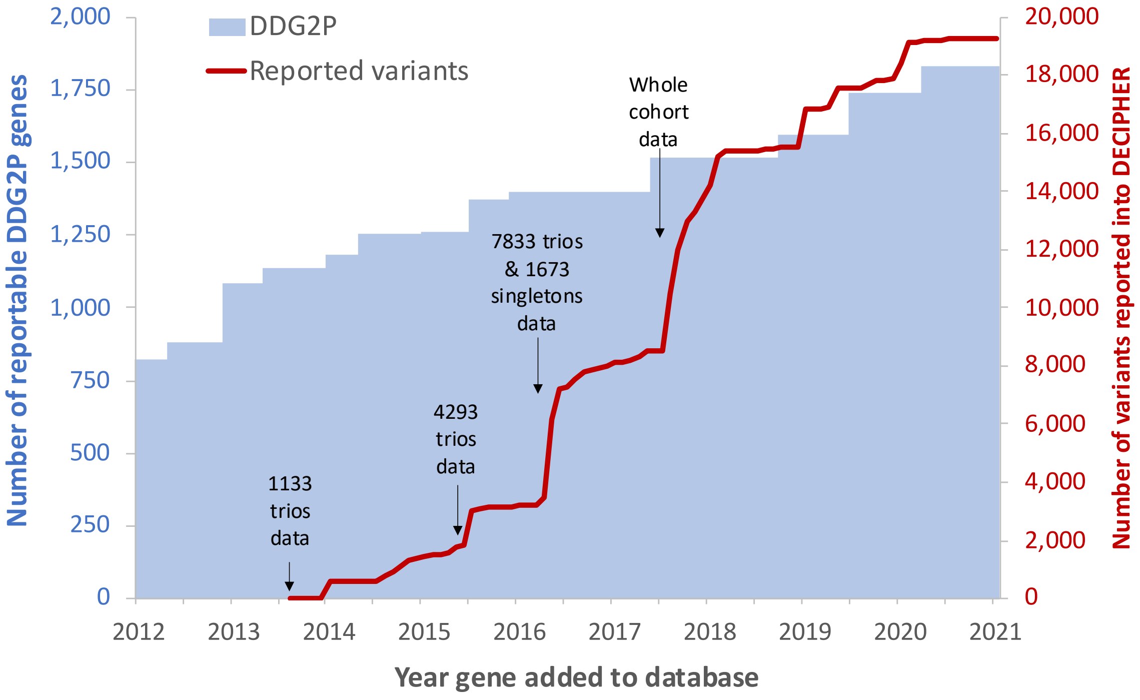

**Figure S2. Candidate diagnostic variants deposited in DECIPHER.**

Sequence variants were detected using WES and included variants <100bp in DDG2P genes; structural variants range from ≥100bp to whole chromosomes and were detected using microarrays and WES.

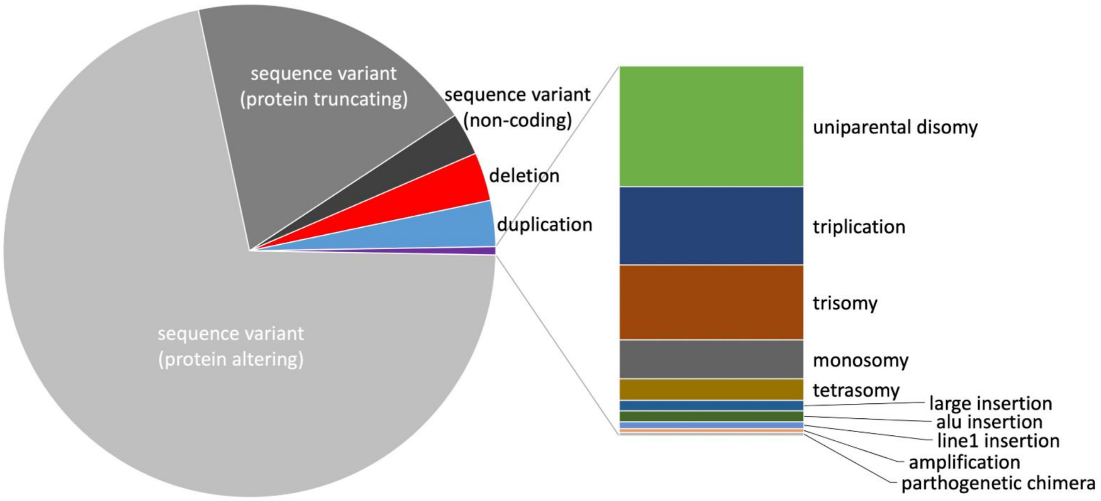

**Figure S3. Number of reported variants per proband.**

Probands are separated into singletons (blue) and trios where neither (yellow), one (orange) or both (red) parents are affected by a shared developmental phenotype. Outliers with >10 reported variants (N=3) were excluded.

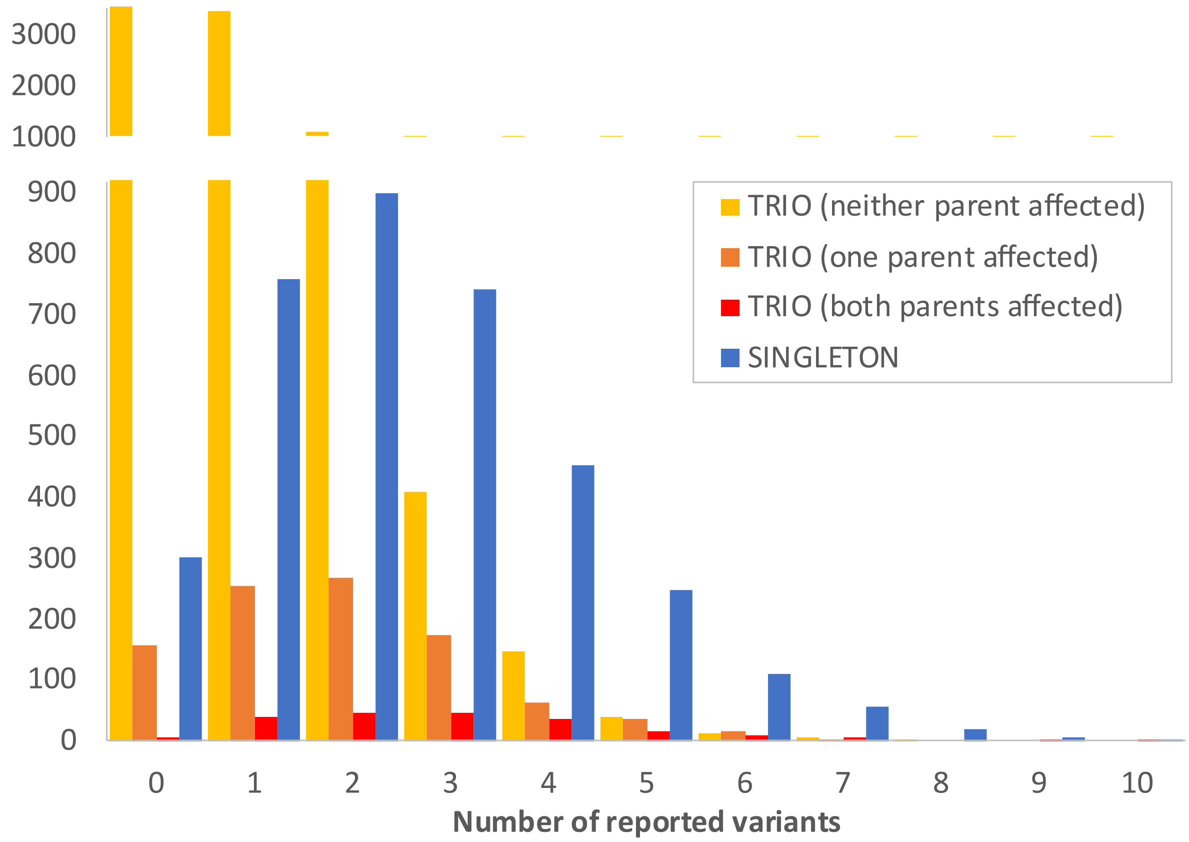

**Figure S4. CADD versus REVEL scores for high impact *de novo* variants.**

*De novo* mutations from 31,058 DD trios[24] were selected to significantly enrich for high impact variants, i.e. absent from gnomAD and functional predicted consequence in a relevant DDG2P gene. Conservative thresholds for applying PP3 (red) and BP4 (green) were then selected.

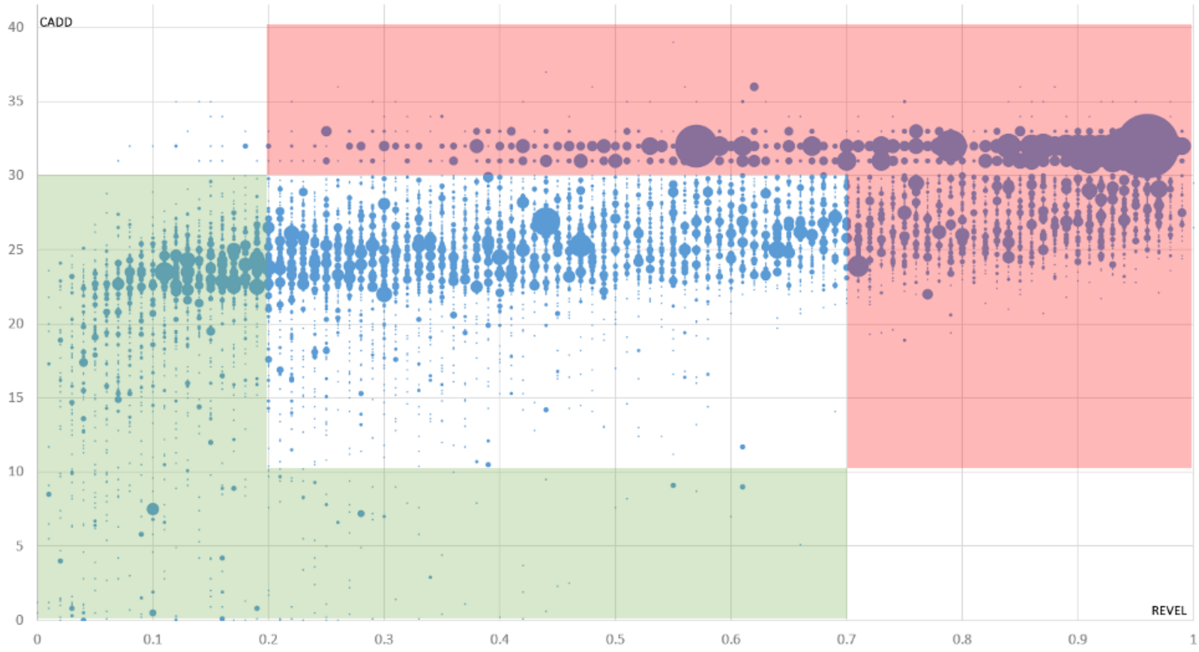

**Figure S5. Concordance between calculated Bayesian posterior probability of variant pathogenicity and clinical classifications.**

Prior probabilities were set to 0.1 for all variants, and ACMG/ACGS variant classification criteria were combined using the log-additive Bayesian likelihood method at very strong, strong, moderate or supporting. Figure created using ggplot2 violinplot (scaled to equal width) in RStudio.

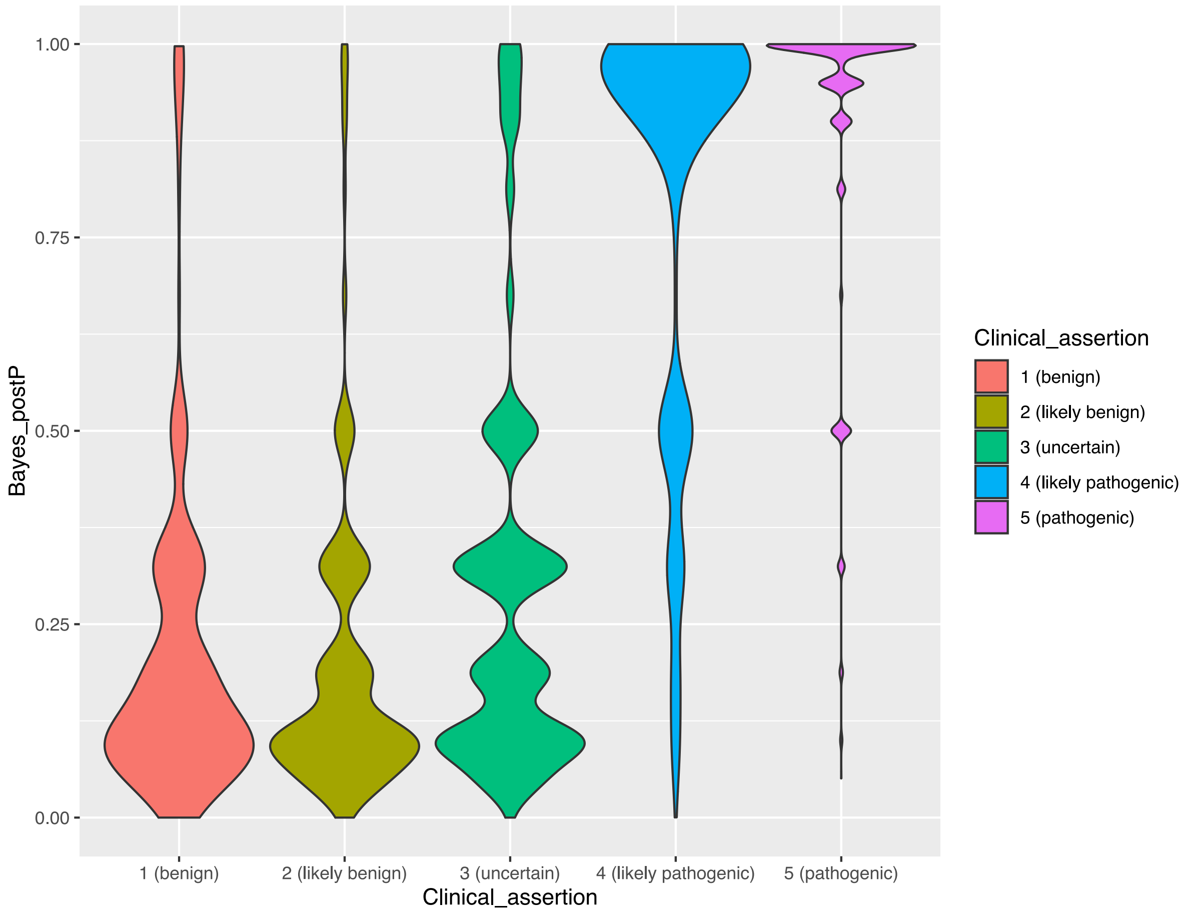

**Figure S6. Affected status of first-degree relatives alters diagnostic rate.**

Odds ratios and 95% confidence intervals of different relative affected statuses in a multivariable regression model controlling for all covariates described in **Table S2** with outcome of diagnosis. Probands without any affected first-degree relatives (N = 10,040) act as the comparison categorical variable. Sibling and parental affected status were determined through standard questionnaires completed by clinical teams via DECIPHER; parental affected status was augmented by collection of relevant/shared HPO terms.

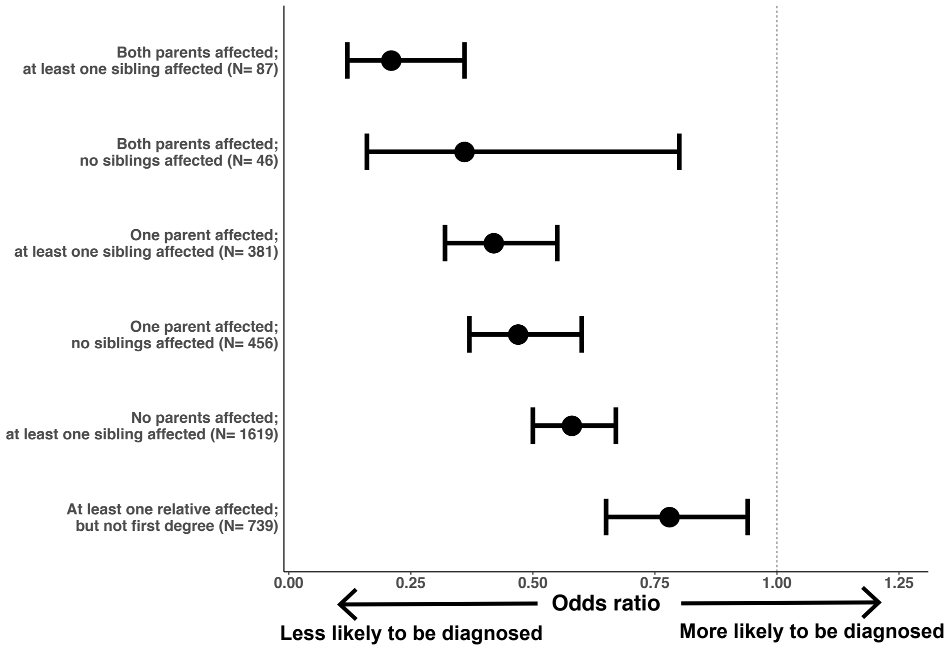

**Figure S7. Comparison of the odds of diagnosis for non-European ancestry probands between trio and singleton cases.**

Odds ratio and 95% confidence intervals for odds of diagnosis in non-European ancestry individuals compared to European-ancestry individuals, controlling for all factors in **Table S2**. The model was applied separately in trios (teal) and singleton (orange) cases with European trios and singletons acting as the baseline in the respective models.

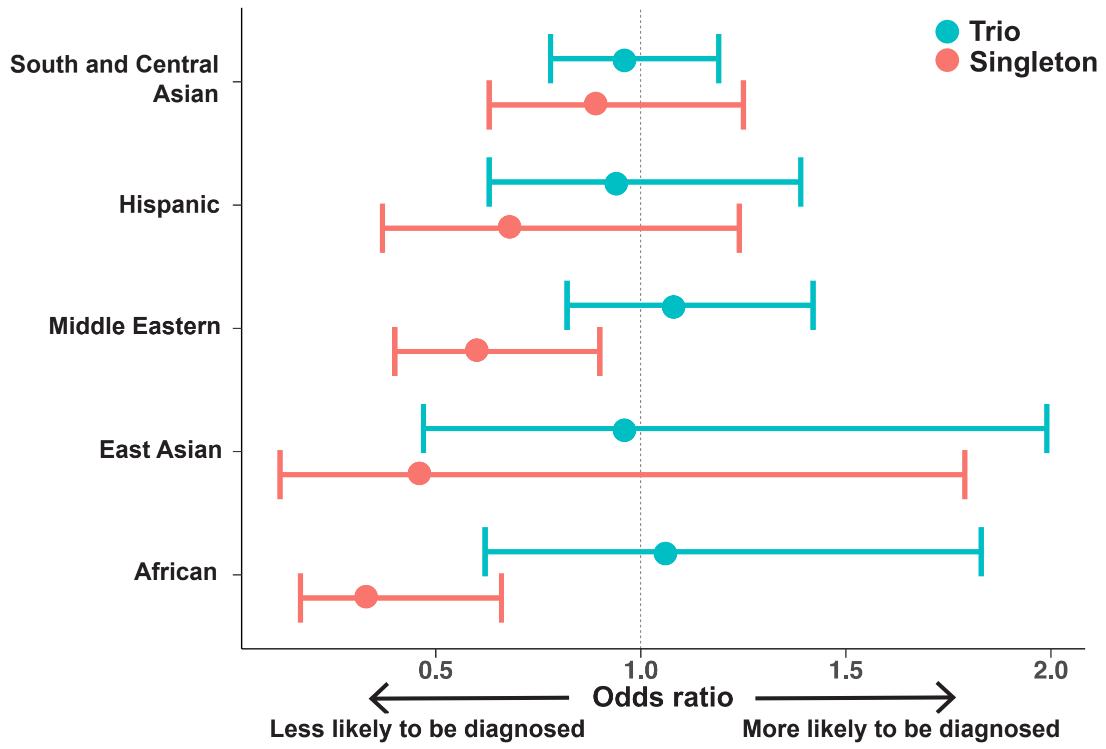

**Figure S8. Breakdown of variants reported via DECIPHER across ancestry groupings.** **(a)** Average number and **(b)** percentage of reported variants per proband, stratified by ancestry group and trio status, with the different colours indicating the average number of variants annotated as Pathogenic/Likely Pathogenic, Benign/Likely Benign, uncertain or as-yet-unrated by clinicians. **(c)** Average number and **(d)** percentage of reported variants separated into European and non-European groupings on a per proband basis. Non-European singletons have significantly more variants reported back on average than European singletons (mean EUR: 2.3; mean non-EUR = 3.3; Wilcoxon p-value = $<2.2\times10^{-16}$). Non-European trios have a smaller increase, but still significant, in the number of variants reported back on average compared to European trios (mean EUR = 0.9; mean non-EUR = 1.1; Wilcoxon p-value = $1.1\times10^{-10}$). Variant pathogenicity is based on clinical assertion only.

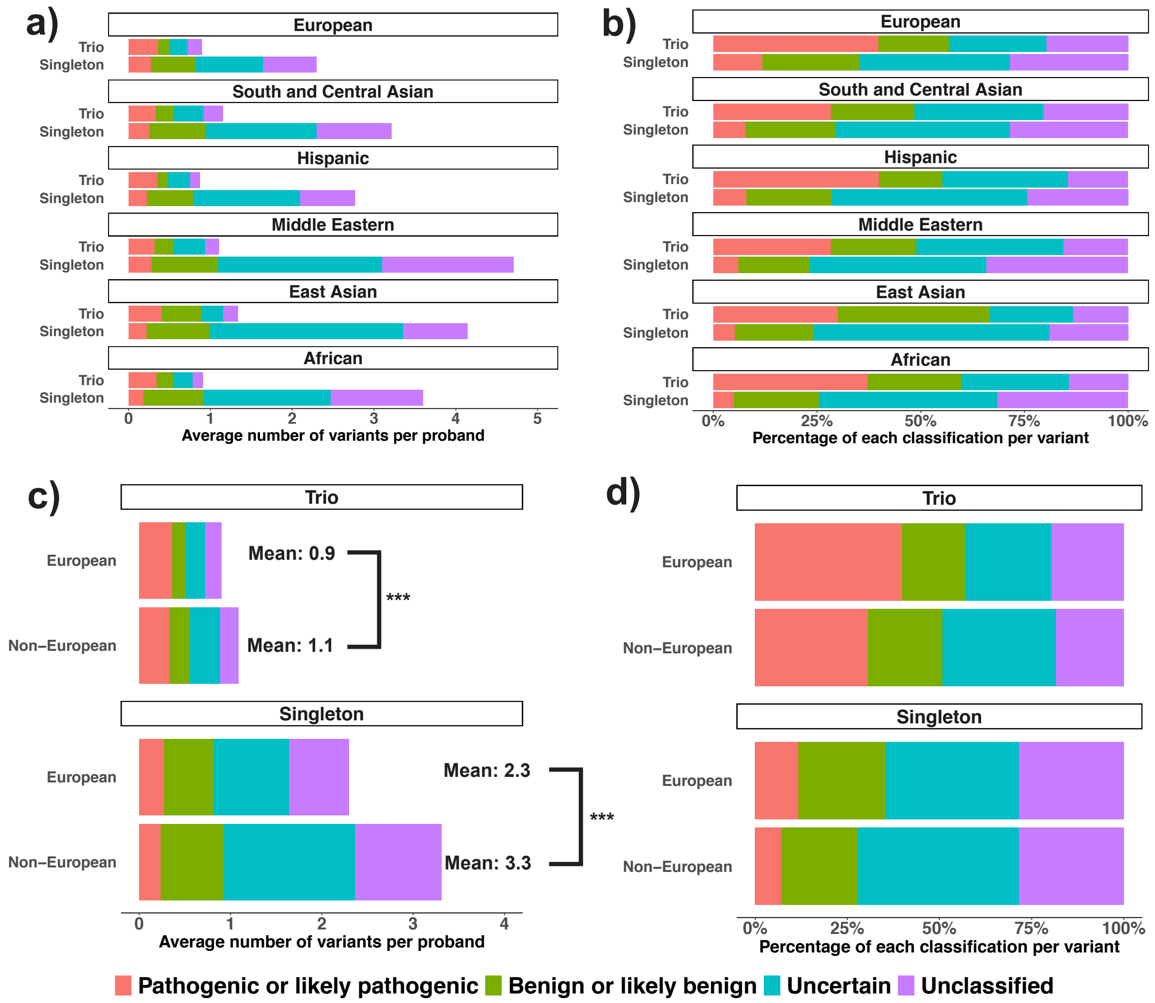

### Supplementary Tables

**Table S1. Proportion of reported variants per variant detection algorithm.**

| **Variant detection algorithms** | **Description** | **Reported variants** |
|---|---|---|
| BWA, GATK, SAMtools | SNVs/indels | 74% |
| DeNovoGear | *De novo* SNVs/indels | 19% |
| CoNVEx, XHMM, CLAMMS, Canoes, CNsolidate | CNVs | 6% |
| Shearwater, DeNovoGear | Mosaic *de novo* SNVs/indels | 0.7% |
| Indelible | Large *de novo* indels | 0.3% |
| MoChA, MAD, triPOD, MrMosaic, chr counter | Large and mosaic structural variants | 0.3% |
| UPDio | Uniparental disomy | 0.2% |
| MELT | Mobile element insertions | 0.05% |

**Table S2. Covariates included in the multivariable logistic regression.**

| | Variable | Number of individuals (Total: 13368) | Derivation |
|---|---|---|---|
| **Binary** | Sex | Male (1): 7818<br>Female (0): 5550 | Chromosomal sex |
| | Trio Status | In trio (1): 9807<br>Not in trio (0): 3561 | Whether the proband was sequenced and analysed as part of a trio |
| | Proband has died | Yes (1): 122<br>No (0): 13246 | Clinical recruitment questionnaire in DECIPHER |
| | Features suggestive of syndrome | Yes (1): 4454<br>No (0): 8914 | Clinical recruitment questionnaire in DECIPHER |
| | Seizure phenotype | Yes (1): 2466<br>No (0): 10902 | Derived from the proband's HPO terms selecting all probands with a descendant term of 'seizure' (HPO: 0001250). This was performed using the ontologyIndex R package version 2.7. |
| | Maternal Diabetes | Yes (1): 482<br>No (0): 12886 | Clinical recruitment questionnaire in DECIPHER |
| | Maternal history of pregnancy loss | Yes (1): 2763<br>No (0): 10605 | Clinical recruitment questionnaire in DECIPHER |
| | Maternal use of antiepileptic medications | Yes (1): 175<br>No (0): 13193 | Clinical recruitment questionnaire in DECIPHER |
| **Categorical** | Ancestry | European (baseline): 11202<br>South & Central Asian: 1252<br>Hispanic: 462<br>Middle Eastern: 243<br>East Asian: 59<br>African: 150 | Genetically inferred ancestry. European ancestry acts as the baseline. |

| | | | |
|---|---|---|---|
| | Recruitment centre | 24 recruiting centres | Clinical recruitment questionnaire in DECIPHER. Centre is not shown but is included in the model. |
| | Severity of Intellectual disability (ID)/ Developmental delay (DD) | None (baseline): 4744<br>Mild: 937<br>Moderate: 1893<br>Severe: 1543<br>Unspecified severity: 4251 | Derived from the proband's HPO terms. Probands with no ID/DD act as the reference. |
| | Neonatal Intensive Care Unit (NICU) admission | Yes: 4067<br>No (baseline): 8627<br>Unknown: 674 | Clinical recruitment questionnaire in DECIPHER. Probands with 'unknown' are not shown in the plots but are present in the model as a categorical group. They were not significantly different from the baseline "No" group. |
| | Only affected member of family | Yes: 9617<br>No (baseline): 3461<br>Unknown: 290 | Clinical recruitment questionnaire in DECIPHER. Probands with 'unknown' are not shown in the plots but are present in the model as a categorical group. They were not significantly different from the baseline "No" group. |
| | Gestation | Extremely premature (22-27wks): 111<br>Very premature (28-31wks): 291<br>Moderately premature (32-36wks): 1744<br>Term (baseline): 11222 | Clinical recruitment questionnaire in DECIPHER: numerical gestation binned into known clinical categorical groupings. Term gestation was treated as the baseline. |
| **Continuous** | Age of proband at recruitment | 13368<br>(mean: 8.20y; SD: 6.37y) | Clinical recruitment questionnaire in DECIPHER. The unit of measurement is years, i.e. the result plotted in Figure 4 is the change in odds of diagnosis for each year increase in proband age |

| | | | |
|---|---|---|---|
| | Time since consent | 13368<br>(mean: 7.44; SD: 1.14) | Clinical recruitment questionnaire in DECIPHER. The unit of measurement is years, i.e. the result plotted in Figure 4 is the change in odds of diagnosis for each year increase in recruitment age. |
| | Number of organ systems affected | 13368<br>(mean: 3.53; SD: 1.79) | Counted as described in Mari et al. (2018).[10] This method avoids double-counting HPO terms that fall under multiple organ systems. The unit of measurement is the number of organ systems affected, i.e. the result plotted in Figure 4 is the change in odds of diagnosis for each additional organ system affected. |
| | Number of Variants in DECIPHER | 13368<br>(mean: 1.34; SD: 1.40) | Downloaded from DECIPHER database on 30 June 2021. Number of variants determined for each proband. Compound heterozygous variants were counted as one variant. The unit of measurement is variants. This is not shown but is included in the model. |
| | Consanguinity | 13368<br>(mean: 0.20; SD: 0.37) | Determined from the fraction of the genome in runs of homozygosity in the genome ($F_{ROH}$). This unit of measurement has then been scaled by 0.0625 (the expected $F_{ROH}$ for the offspring of first cousins) to make the odds more interpretable, i.e the result plotted in figure 4 is the change in odds of diagnosis for each increase in autozygosity equivalent to the offspring of first cousins. |

**Table S3. Trio information across different ancestry groupings.**

Total number of probands in the study from each genetically defined ancestry group, the number in trios and the percentage in trios. Also included are the p-values for interaction between trio and ancestry from in the multivariable logistic regression. The European group acts as the baseline to which other ancestries are compared.

| Ancestry | Total probands | Probands in trios | Percentage in trios | Interaction Trio*Ancestry (P-Value) |
|---|---|---|---|---|
| **European** | 11202 | 8294 | 74% | NA |
| **South and Central Asian** | 1252 | 915 | 73% | $6.9 \times 10^{-3}$ |
| **Hispanic** | 462 | 297 | 64% | $7.5 \times 10^{-4}$ |
| **Middle Eastern** | 243 | 179 | 74% | $4.9 \times 10^{-4}$ |
| **East Asian** | 59 | 45 | 76% | 0.05 |
| **African** | 150 | 77 | 51% | $1.7 \times 10^{-5}$ |

16. King DA, Sifrim A, Fitzgerald TW, et al. Detection of structural mosaicism from

targeted and whole-genome sequencing data. Genome Res 2017;27(10):1704–14.

30. Ellard S, Baple E, Callaway A, Berry I, Forrester N, Clare. ACGS Best Practice Guidelines for Variant Classification in Rare Disease 2020. 2020;
